# Whole genome sequencing reveals a decade of human adenovirus B114 circulation in England

**DOI:** 10.64898/2026.09.22.26363659

**Authors:** C. Patrick McClure, Maria Muntean, Eva Cifelli, Oladimeji John Abiodun, Michelle M. Lister, Arnold W. Lambisia, Joshua Quick, Stuart Astbury, Charlotte J. Houldcroft

## Abstract

**Background:** Human Mastadenovirus B (HAdV-B) causes ocular and respiratory infections and is capable of causing outbreaks in healthcare settings. HAdV-B114, a recombinant genotype only formally recognised in 2023, has been reported predominantly from Germany, the USA, China and Japan; genomic data from UK clinical settings remain limited. This study analysed 68 HAdV-B whole genomes from ocular and respiratory-associated samples collected at a Nottingham hospital across the pre- and post-COVID-19 restriction periods (2017-2023).

**Methods:** Clinical samples typed positive for HAdV-B by partial hexon Sanger sequencing were selected for whole-genome sequencing using a novel overlapping PCR amplicon strategy, sequenced on the Oxford Nanopore Technologies PromethION platform. Phylogenetic and pairwise difference analyses were used to characterise genotype composition and genomic relatedness. SimPlot and amino-acid comparison characterised divergence between sub-lineages.

**Results:** Whole-genome phylogenetics confirmed that two species B genotypes were detected in Nottingham University Hospitals between 2017-23: HAdV-B114 (n=56) and HAdV-B7 (n=12). HAdV-B114 was detected before and after COVID-19 restrictions, with an increase in 2022-2023, consistent with post-restriction expansion of a pre-existing lineage. Pairwise difference analysis identified 24 genomically identical B114 sequences; associated metadata argued against a single nosocomial transmission chain interpretation, with the pattern more consistent with repeated detection of a widely circulating community strain, with the possibility of localised onward transmission. Within the B114 clade, multiple monophyletic groups were identified by whole-genome phylogenetics.

**Conclusions:** This study adds whole-genome resolution to the existing UK HAdV-B114 data, demonstrates the utility of WGS for clinical adenovirus surveillance and shows the benefits of combining epidemiological data with genomic analysis.

## Introduction

Adenovirus-associated ocular and respiratory infections generate substantial global healthcare burden [1], affecting patients across all age groups and with the capacity to cause outbreaks in healthcare settings. Effective surveillance of circulating strains is therefore an important public health priority, particularly in clinical environments where patient-to-patient transmission can occur.

Whole-genome sequencing (WGS) has transformed viral surveillance by enabling accurate genotype assignment, detection of recombinant strains, assessment of genomic relatedness and identification of lineage structure. This is particularly important for double-stranded DNA human adenoviruses (HAdV), which are classified into seven species (A-G) and evolve substantially through homologous recombination between capsid genes; typing based on a single gene or short genomic region can therefore misclassify recombinant strains and fail to detect diversity elsewhere in the genome [2, 3]. The three major capsid proteins – penton, hexon and fiber – mediate receptor attachment, cell entry and host immune recognition respectively [4], and are the primary basis for adenovirus genotyping and the regions most likely to be subject to adaptive immune pressure.

Despite the growing application of WGS to adenovirus surveillance, UK genomic data for HAdV species B remains limited. Most published whole-genome data for HAdV-B114 – a recombinant genotype only formally recognised in 2023, and previously circulating undetected under misclassification as HAdV-B3 – originate from Germany, the USA, China and Japan [5]. How B114 and related species B genotypes have circulated in UK clinical settings, particularly across the epidemiological disruption of the COVID-19 pandemic, remains poorly characterised.

This study addresses that gap by analysing 68 HAdV-B whole genomes from ocular and respiratory-associated samples collected in a Nottingham hospital, spanning the pre- and post-COVID restriction periods. This represents the largest UK whole-genome dataset for HAdV-B, and one of only two available for HAdV-B114 specifically. This study aimed to confirm circulating genotype composition and describe genotype distribution across the study period; assess genomic relatedness between Nottingham genomes and evaluate whether sequence similarity was compatible with recent patient-to-patient transmission; and characterise sub-lineage structure within HAdV-B114 and identify candidate genomic regions contributing to divergence between sub-lineages.

## Methods

### Ethics and infection control

The study utilized anonymised surplus nucleic acid extracted in routine diagnostic investigation of suspected viral illness at Nottingham University Hospitals National Health Service Trust, who approved extended targeted viral genomic investigation of diagnosed adenoviral positives under clinical audit number 23-078C.

### Samples

Sixty-eight specimens from 66 unique patients were sampled between October 2017 and July 2023 at NUH NHST at various in-and outpatient clinics. Samples were collected across pre-and post-COVID-19 restriction periods (2017-2023) and included ocular and respiratory-associated specimens. Associated metadata included sample type, sample collection week, patient age group and clinic source (Table 1).

**Table 1:** Metadata for the 24 Nottingham HAdV-B114 genomes sharing identical consensus sequences.

| Name | Age group (years) | Week number | Year | Sample Type | Source |
| --- | --- | --- | --- | --- | --- |
| HAdVb114_UKN22_1 | 15-44 | 22 | 2022 | Swab from eye | AAU1 |
| HAdVb114_UKN22_2 | 65-79 | 26 | 2022 | Swab from eye | AAU1 |
| HAdVb114_UKN22_4 | 15-44 | 27 | 2022 | Swab from eye | AAU1 |
| HAdVb114_UKN22_5 | 0-4 | 28 | 2022 | Swab from throat | PMA |
| HAdVb114_UKN22_8 | 65-79 | 30 | 2022 | Swab from eye | AAU1 |
| HAdVb114_UKN22_10 | 0-4 | 31 | 2022 | Nose and throat swab | PMW1 |
| HAdVb114_UKN22_12 | 15-44 | 32 | 2022 | Swab | AAU1 |
| HAdVb114_UKN22_15 | 15-44 | 34 | 2022 | Swab from eye | AAU1 |
| HAdVb114_UKN22_16 | 0-4 | 34 | 2022 | Nasopharyngeal aspirate | PMW2 |
| HAdVb114_UKN22_19 | 0-4 | 38 | 2022 | Swab from eye | OU1 |
| HAdVb114_UKN22_22 | 45-64 | 42 | 2022 | Swab | AAU1 |
| HAdVb114_UKN22_23 | 5-14 | 49 | 2022 | Swab from nose | PMW3 |
| HAdVb114_UKN22_24 | 0-4 | 51 | 2022 | Swab from eye | PMA |
| HAdVb114_UKN22_25 | 15-44 | 53 | 2022 | Swab from eye | AAU1 |
| HAdVb114_UKN22_27 | 5-14 | 53 | 2022 | Swab from nose | PMA |
| HAdVb114_UKN22_26 | 15-44 | 53 | 2022 | Swab from eye | OU1 |
| HAdVb114_UKN23_3 | 15-44 | 2 | 2023 | Swab from eye | AAU1 |
| HAdVb114_UKN23_4 | 15-44 | 3 | 2023 | Swab from eye | AAU1 |
| HAdVb114_UKN23_8 | 15-44 | 7 | 2023 | Swab from eye | AAU1 |
| HAdVb114_UKN23_9 | 0-4 | 7 | 2023 | Swab from throat | PMA |
| HAdVb114_UKN23_10 | 0-4 | 10 | 2023 | Swab | GPS |
| HAdVb114_UKN23_12 | 15-44 | 12 | 2023 | Swab from eye | OU1 |
| HAdVb114_UKN23_14 | 0-4 | 14 | 2023 | Swab from throat | PMA |
| HAdVb114_UKN23_17 | 15-44 | 18 | 2023 | Swab from throat | CC1 |

Total nucleic acid (TNA) extracted (bioMérieux NucliSENS easyMAG system) and eluted in a 50⍰µl volume. Extracts were then diagnosed Adenoviral-positive by either the AusDiagnostics Respiratory 16-plex panel (REF 20602, AusDiagnostics, Australia) or Meningitis / Encephalitis viral 8-plex panel (REF 27093, AusDiagnostics, Australia) in the case of ocular swabs, returning a semi-quantitative output of arbitrary copies per 10ul nucleic acid extract. Surplus adenoviral-positive extracts were initially genotyped by Sanger sequencing of the hexon gene using primers and conditions from Wong et al [6] (forward: GCCACCTTYTTCCCCATGGC; reverse GTAGCGTTRCCGGCNGAGAA). Whole-genome sequences were subsequently generated using an overlapping PCR amplicon scheme sequenced on the Oxford Nanopore Technologies Promethion platform at Deep Seq Nottingham as previously described [7].

### Sequencing method

#### Overlapping amplicon design and QC

Primers were designed using Primalscheme [8] and downloaded from https://github.com/quick-lab/HAdV/tree/main/HAdV-B/v1.0 to generate an overlapping scheme of 16 amplicons ranging in size from 2051 to 2897bp (as described in [9]). Synthesized primers were resuspended at 100uM and mixed in two equimolar pools for either odd or even numbered amplicons, with the exception of those for amplicon targets 2, 4, 6, 8, 10, 11, 15 and 16 where double volume was added after initial universally equimolar testing. Primers pools were diluted 1 in 10 and 1ul of each pool was used in separate multiplex PCR reactions as previously described. Briefly, two 20ul reactions were assembled with Q5 2x Hot Start Master Mix (NEB) containing 1ul of primers targeting either odd or even numbered amplicons and 2ul nucleic acid extract.

#### Sequencing method and QC

Sequencing libraries were prepared using the Oxford Nanopore Native Barcoding kit (SQKNBD114.96) and sequenced on R10.4.1 flowcells on a PromethION sequencer. Basecalling was carried out using Dorado (v2.1.2) using the high accuracy model (v4.3.0).

#### Consensus calling and mapping

Pass reads (average Phred score ≥7) were used as the input for artic guppyplex to carry out length filtering based on the amplicon scheme (2000-3000bp). These filtered reads were then used as the input for the ARTIC fieldbioinformatics pipeline (v.1.8.5, https://github.com/artic-network/fieldbioinformatics). Within this pipeline reads were aligned to reference sequences and corresponding primer .bed files based on appropriate subtype (B114: OR853835, B7: MH921836), and Clair3 (v1.2.0) was used to call variants against these reference sequences. Before variant calling all alignments were normalised to 200x depth, and remaining settings were left as defaults with any site dropping below 20x coverage replaced with an “N”.

### Sequence alignment and phylogenetic analysis

Complete human Mastadenovirus B genomes from genotypes 3, 7, 114 and known or possible recombinants were downloaded from GenBank on 01/05/2026. Several sequences labelled as human mastadenovirus B were manually removed after nucleotide blast analysis identified them as more similar to species human mastadenovirus C. Duplicate sequences, sequences derived from patents or labelled Modified Microbial Nucleic Acid were also removed. Sequences with greater than 1% Ns were removed. The dataset includes pre-2020 HAdV-B genomes from Great Ormond Street Hospital (GOSH) in London, UK (Myers et al.,2021).

Filtered sequences were aligned to the B3 RefSeq NC_011203 using MAFFT online, with the keep-length and treat N as wildcard options [10]. The total dataset was split into two in order to improve the computational running time of whole-genome phylogenetic analyses:

1. B7 + B7 recombinants + UK adenovirus B sequences
2. B3 + B114 + B3/B114 recombinants + UK adenovirus B sequences

The final B7 alignment included 290 sequences of length 35343 bases. The final B3/B114 alignment included 235 sequences of length 35343 bases.

Maximum-likelihood tree was constructed using raxmlGUI 2.0 [11], with the substitution model chosen using model test in raxmlGUI. The best tree was determined through 100 bootstrap analyses using a GTR+Gamma model. Trees were visualised in iToL [12] and clades which did not contain reference sequences or samples from the UK were collapsed. Bootstrap values over 70 are shown.

A maximum likelihood tree was then made as before with all Nottingham (including samples with greater than 1% Ns, identical sequences and paired patient samples) and UK B3, B7, B11 and B114 samples in Genbank, constructed in raxml2.0 GUI and visualised in iToL. Additional Genbank sequences manually identified as closely related were added as international context, giving a total of 188 sequences.

### Pairwise difference analysis

Pairwise whole-genome nucleotide differences between Nottingham sequences were calculated in MEGA v12. Pairs of sequences differing by 0-3 single nucleotide polymorphisms (SNPs) were considered compatible with very recent common ancestry (reference) and treated as candidate transmission-related pairs; given the slow rate of sequence change in double-stranded DNA viruses, a threshold of 0-3 SNPs is consistent with transmission within a short epidemiological window. However, genomic similarity alone was not taken as sufficient evidence of direct patient-to-patient transmission.

### SimPlot analysis and amino-acid comparison

Following whole-genome phylogenetic analysis, two sub-lineages were identified within the HAdV-B114 clade. One representative complete genome from each sub-lineage was selected for detailed comparison: HAdVB114_UKN22_7 (sub-lineage A) and HAdVB114_UKN22_23(sub-lineage B).

SimPlot++[13] was used to visualise genome-wide nucleotide similarity and identify the distribution of differences between representatives (window size 200 bp; step size 20 bp).The HAdV-B114 prototype genome (OR853835.1) was used for comparison.

### Statistical tests

The distribution of case-ages by genotype were tested for normality with the Lilliefors test, and were found not to be normally distributed. Therefore a non-parametric test (Kruskal-Wallis) of the difference in age distributions was carried out. For individuals with more than one sample, only the first sample was included in this analysis. Statistical tests were carried out in Matlab 2025b.

## Results

### Amplicon sequencing successfully recovers whole adenovirus genomes from archive clinical samples

The priming scheme was initially evaluated on 4 samples, ranging from approximately 1000 to 175,000 genome equivalents per reaction. Whilst only amplicon 2 dropped below the minimum 20x coverage threshold in the two lower titre samples, sub-optimal depth (<190x) was observed in several other amplicons. A final version of the scheme with primers targeting low-depth amplicons doubled in concentration was applied to a subset of forty samples ranging in viral titre from 13 to 1229855 genome equivalents, achieving thirty seven complete genomes and only single amplicon dropout in the remaining three. Two of these were in amplicon 8, with relatively lower template inputs of <250 copies and one in amplicon 16 respectively with a comparatively high viral titre unput of 41825 copies. No primer mismatch was identified as a cause of amplicon dropout, although this was not possible to assess in the case of the reverse amplicon 16 primer for HAdVb114_UKN22_16.

### Whole-genome phylogeny confirmed circulating genotypes

Whole-genome phylogenetic analysis grouped Nottingham sequences into two genotypes: HAdV-B114 (n=56) and HAdV-B7 (n=12). HAdV-B sequences from GOSH (London, UK) provided a pre-2020 UK reference point (Figure 1). UKHSA B7 genomes collected from England post-2020 were also available. B7 and B114 samples fell in distinct clades, with no apparent recombinants between Nottingham genotypes.

**Figure 1:**
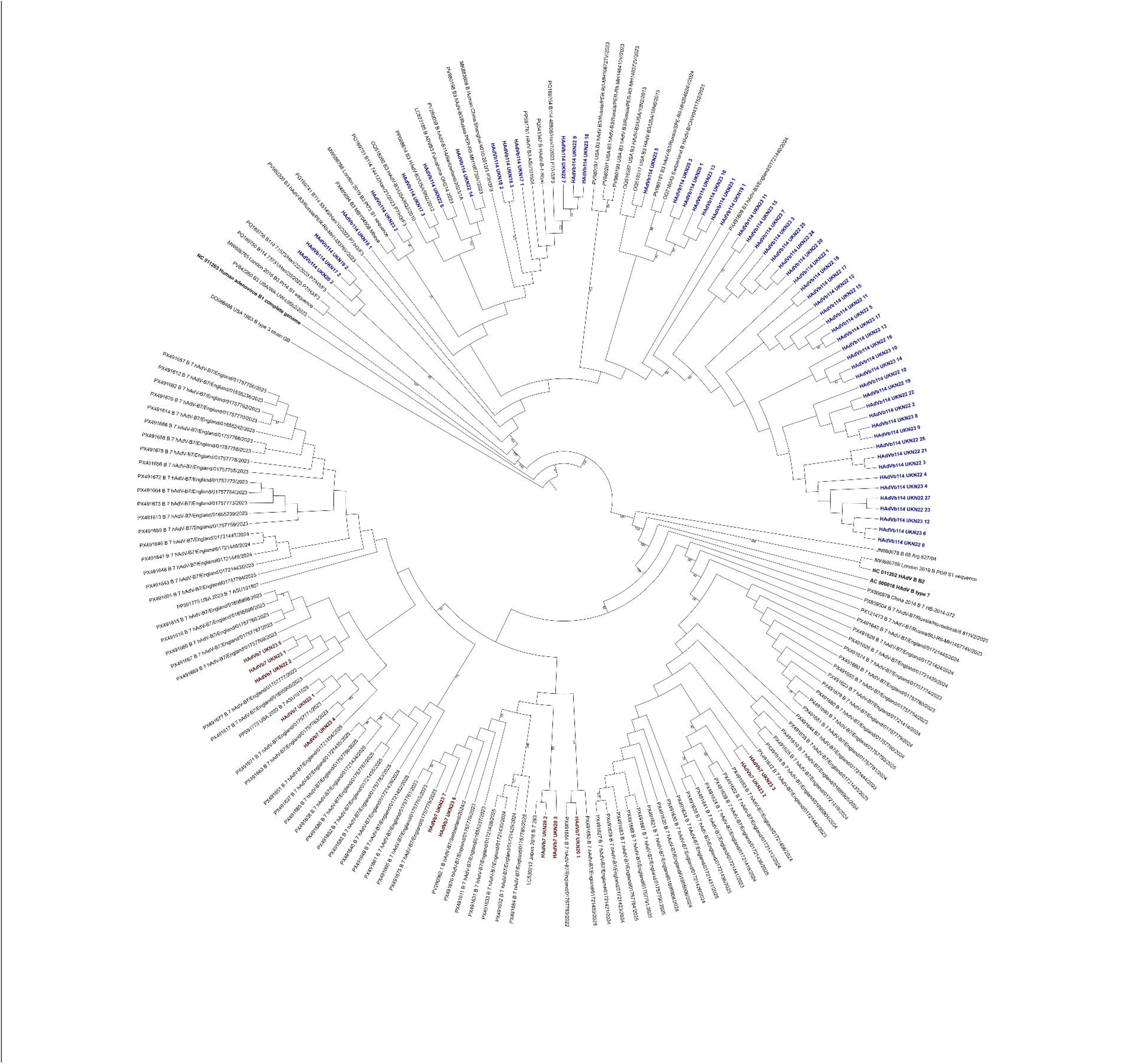
Whole genome maximum likelihood phylogenetic tree of 68 Nottingham HAdV-B sequences, HAdV-B reference genomes for types B3, B7 and B11 (shown in bold black text), and UK adenovirus B3, B7, B11 and B114 sequences recovered from GenBank. Bootstrap values over 70 (100 replicates) are shown at each node. Samples identified by blast analysis and phylogenetic placement as B114 are shown in blue; samples identified as B7 are shown in dark red. Tree constructed in RAML2.0 GUI and visualised Using Itol web

We then placed the unique adenovirus B sequences from Nottingham in global phylogenetic context (Figure 2). Figure 2A shows that the Nottingham B114 sequences are highly similar to, and cluster phylogenetically with, B114 genomes sequenced from China, the USA, Germany, England, Russia and Japan. The closest phylogenetic relatives of Nottingham B7 genomes (Figure 2B) come from England, Switzerland, and Japan. These data are likely to reflect the countries performing adenovirus genomic surveillance, rather than implying direct transmission.

**Figure 2:**
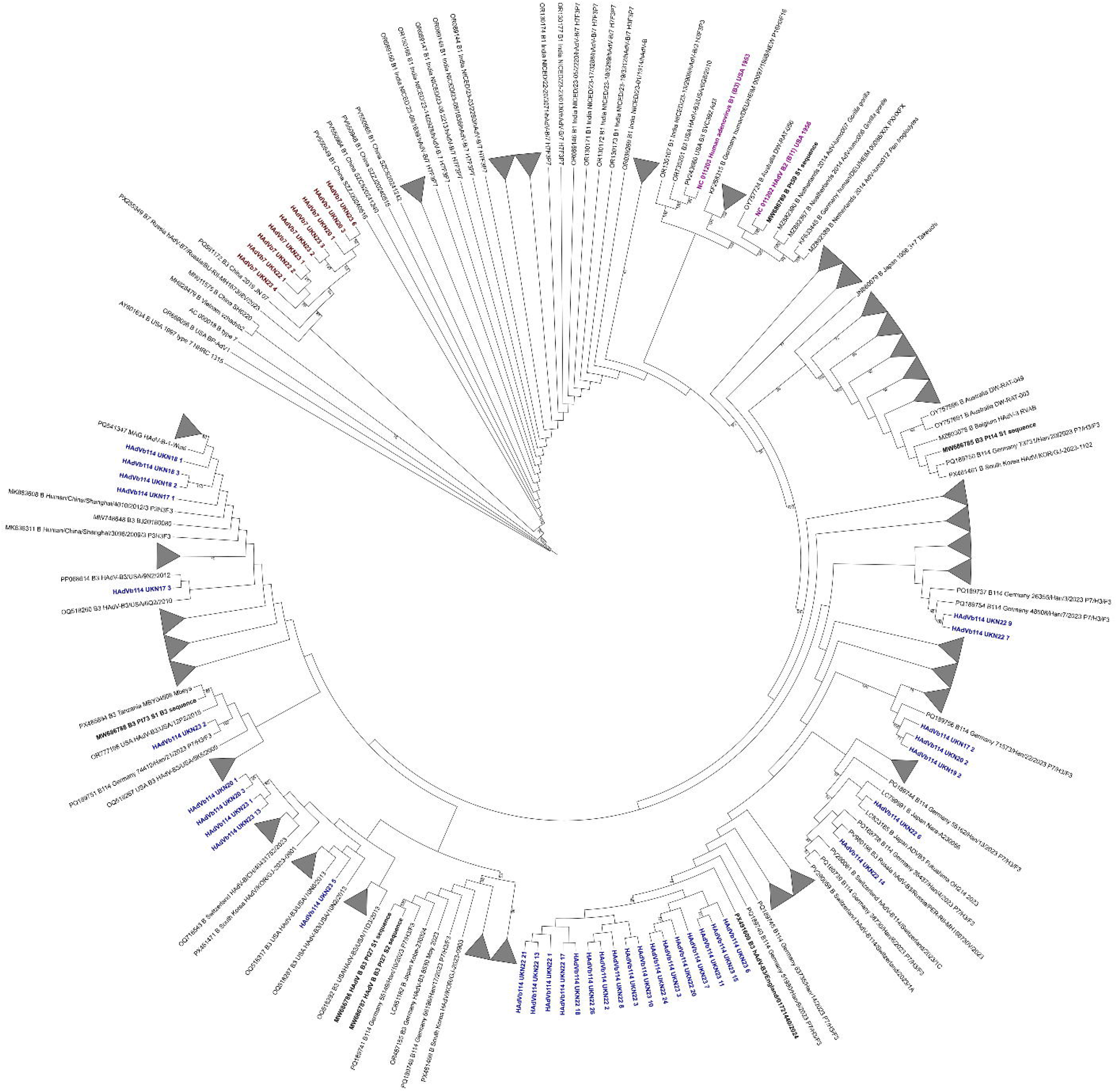
A. Whole genome maximum likelihood phylogenetic tree of 68 Nottingham HAdV-B sequences, HAdV-B reference genomes for types B3, B7 and B11 (shown in bold purple text), and B3 and B114 sequences from GenBank. UK samples are shown in bold. Bootstrap values over 70 (100 replicates) are shown at each node. Clades containing no Nottingham samples were collapsed. B114 genomes from Nottingham are shown in blue; B7 genomes are shown in dark red. Tree constructed in RAXML2.0 GUI and visualised using Itol web. B. Whole genome maximum likelihood phylogenetic tree of 68 Nottingham HAdV-B sequences, HAdV-B reference genomes (shown in bold purple text), and B7 sequences from GenBank.

### HAdV-B114 was detected before and after COVID-19 restrictions

To assess whether HAdV-B114 was present in the Nottingham dataset prior to COVID-19 restrictions, genotype counts were plotted by year of sample collection. A small number of HAdV-B114 genomes were identified from the pre-pandemic period, establishing that this genotype was circulating in Nottingham before COVID-19 restrictions were introduced. Three sequences from two patients (MW686785-MW686787), collected in London in 2015, were initially genotyped as B3 [14] but cluster with genotype B114 samples from the UK and Germany. Few genotyped samples were available in Nottingham in 2020 and none in 2021. An increase in B114 genomes was observed in 2022-23 (Figure 3A). Both B114 and B7 were identified across a broad age range (Figure 3B); the mean age of B7-associated cases (15.9 years) was lower than that of B114-associated cases (20.7 years). The age distributions of B7 and B114 cases were not significantly different (Kruskal-Wallis test, p = 0.74, a = 0.01).

**Figure 3:**
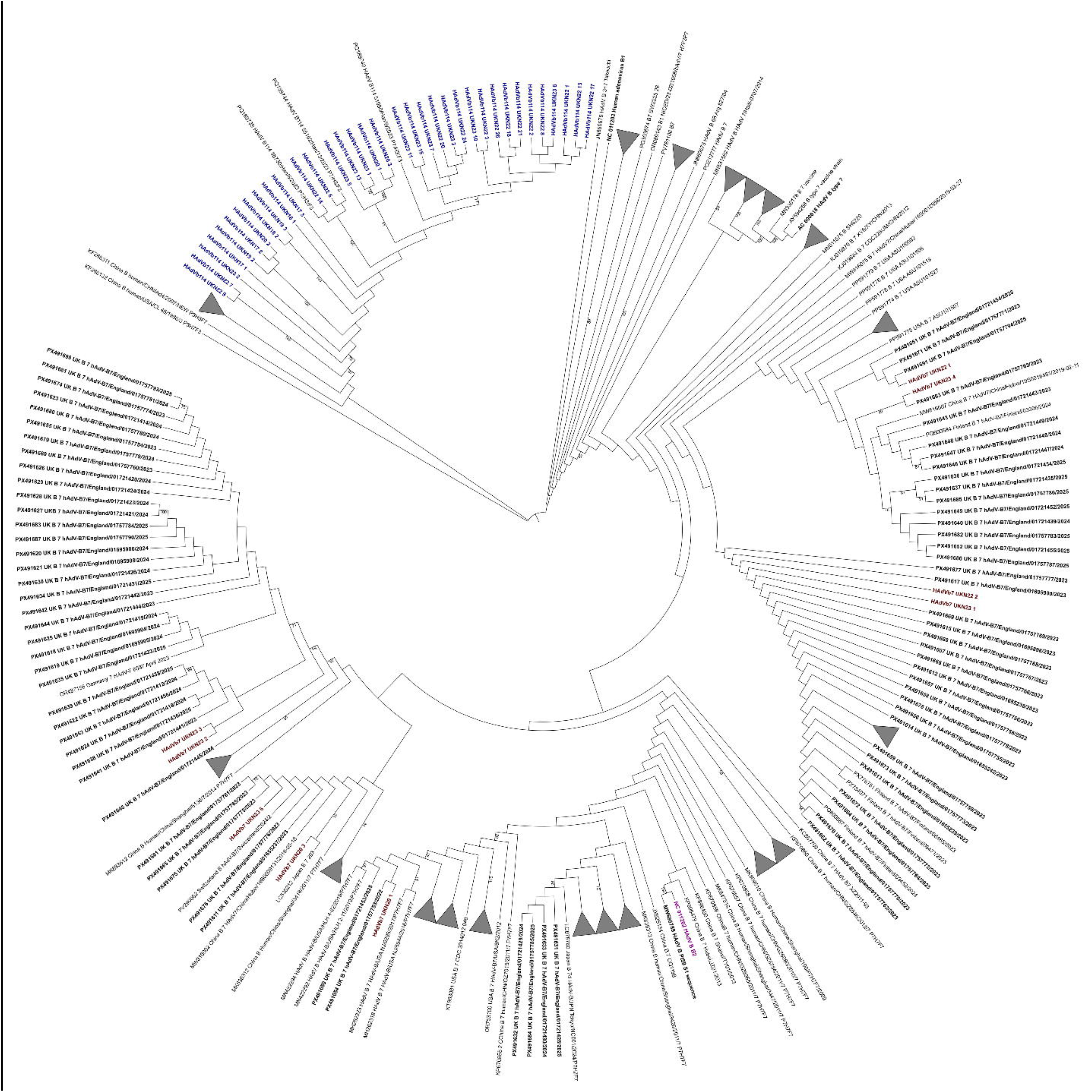
A. Overview of HAdV-B genotypes detected in Nottingham samples (2017–2023). Stacked bars show the yearly number of WGS-typed specimens assigned to HAdV-B7 (blue) and HAdV-B114 (orange). No specimens were genotyped in 2021. B. Age distribution of patients infected with HAdV-B7 and HAdV-B114 in the Nottingham cohort. Box plots show the median (horizontal line), interquartile range (box), and full data range (whiskers).

### Human adenovirus B7 was independently introduced into the Nottingham patient population multiple times between 2020 and 2023

Using a pairwise SNP cutoff of 0-3 SNPs between samples to define a potential transmission cluster, we identified three clusters of B7 sequences which met this criterion. The largest group, group 3, contained five members (HAdVb7_UKN22_1, HAdVb7_UKN22_2, HAdVb7_UKN23_1, HAdVb7_UKN23_5, and HAdVb7_UKN23_4). Phylogenetic analysis of all Nottingham and UK B7 genomes in Genbank supported the identification of three monophyletic groups of Nottingham B7 samples. Samples in groups 1 and 2 were sampled in 2023, while samples in group 3 were collected between 2022-2023. These groups are similar to sequences from elsewhere in the UK (group 1), UK and Switzerland (group 2), and UK and USA (group 3). A sample collected in the USA (PP591775), which was collected in Arizona in March 2023, only differed from HAdVb7_UKN22_1 by a single pairwise difference despite collection dates that are months apart. Combining these data with the temporal range over which samples were collected argues against direct patient to patient transmission; we hypothesise that these represent multiple introductions of HAdV B7 into the Nottingham patient population between 2020 and 2023.

### A large cluster of genomically identical genomes was identified in outpatients attending emergency assessment clinics identified within the Nottingham patient population

To assess genomic relatedness and identify clusters of closely related genomes in this Nottingham dataset, pairwise difference analysis was performed. This revealed that 24 HAdV-B114 genomes were identical at the whole-genome consensus level (zero pairwise SNP differences) (group 9, Figure 4B). The 24 genomes spanned approximately eleven months of sample collection, from May 2022 to May 2023, encompassed a broad age range (3 months to 68 years), included both ocular and respiratory specimens and originated from multiple distinct clinic sources (Table 1). Within the cluster, several sub-groups of samples shared both clinic source and short collection windows. However, one UK sample from Genbank, PX491609, with a collection date of December 2024, differs from many of the Nottingham group 9 samples by a single pairwise difference.

**Figure 4:**
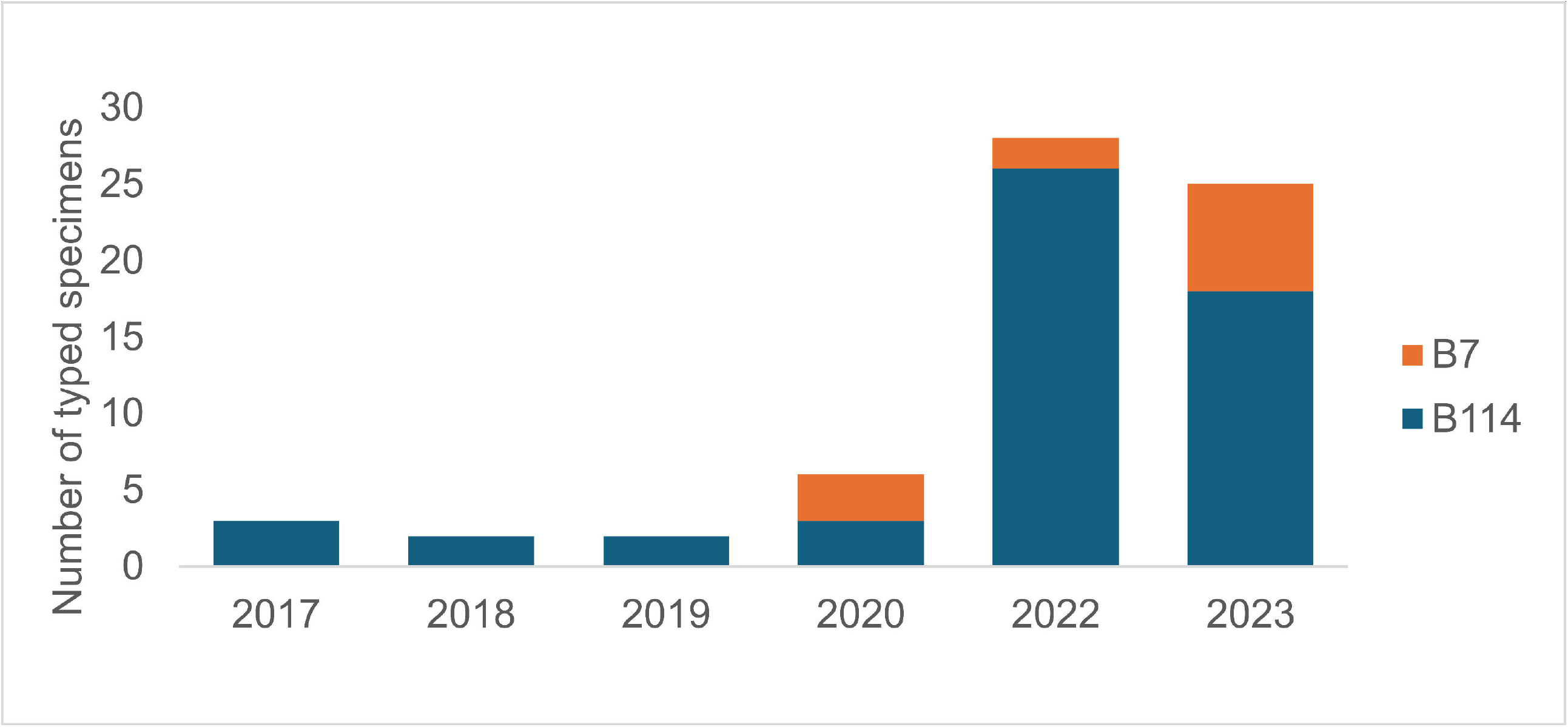
Maximum likelihood phylogenetic tree of B7 (A) and B3 and B114 sequences from this study. Trees were visualised in iTol [12]. Groups are defined as monophyletic clusters containing pairs of sequences with ≤ 3 pairwise differences. **A** shows all B7 sequences from this study; the clade containing Nottingham B114 sequences is collapsed. **B** shows all B3 and B114 sequences from this study; the clade containing Nottingham B7 sequences is collapsed.

The combination of in-patient, out-patient and community samples, plus English public health samples of unknown sampling location, suggests that cluster 9 was the not the result of nosocomial transmission but rather a regional or national epidemic of B114 ‘spilling in’ to the Nottingham patient population.

Clinic source abbreviations: AAU1 Acute Assessment Unit 1, CC1 Community Clinic 1, GPS a general practice surgery from a northerly suburb of Nottingham, OU1 Outpatients Unit 1, PMA Paediatric Medical Assessment, PMW1 Paediatric Medical Ward 1, PMW2 Paediatric Medical Ward 2, PMW3 Paediatric Medical Ward 3.

Beyond the main cluster of 24 identical genomes and further 10 genomes with 1-3 pairwise differences, several smaller groups of identical or near identical sequences were also identified, including three additional B114 clusters (groups 4-8) (Figure 4B). These groups are often closely phylogenetically related to samples from other countries.

Group 6 contained prepandemic samples, groups 4, 8 and 9 contained both pre-and post-pandemic samples, while groups 5 and 7 were composed of of post-restriction genomes. The presence of multipe clusters, some spanning multiple years, indicates that B114 diversity in the Nottingham dataset was not simply a single uniform lineage.

### Divergence between groups was limited but included a difference at hexon codon 204

We next asked whether whether post-pandemic group 7 and pre-/post-pandemic group 9 differed in biologically important genomic regions. To quantify and localise the genomic differences between the B114 groups, one representative genome from each cluster was selected for detailed comparison HAdVB114_UKN22_23 (group 9) and HAdVB114_UKN22_7 (group 7), with a prototype B114 sequence from Gemany (OR853835) used as the comparator strain in SimPlot. Pairwise difference analysis identified 16 SNPs between the two Nottingham group 7 and 9 representatives across the whole genome. SimPlot was then used to visualise the genomic distribution of the 16 differences (Figure 5). The similarity score remained at approximately 0.995 – 1.000 throughout, confirming that the two sub-lineages were closely related rather than deeply divergent. The distribution of SNPs across the genome suggests sequence evolution driven by nucleotide mutations rather than recombination [15].

**Figure 5:**
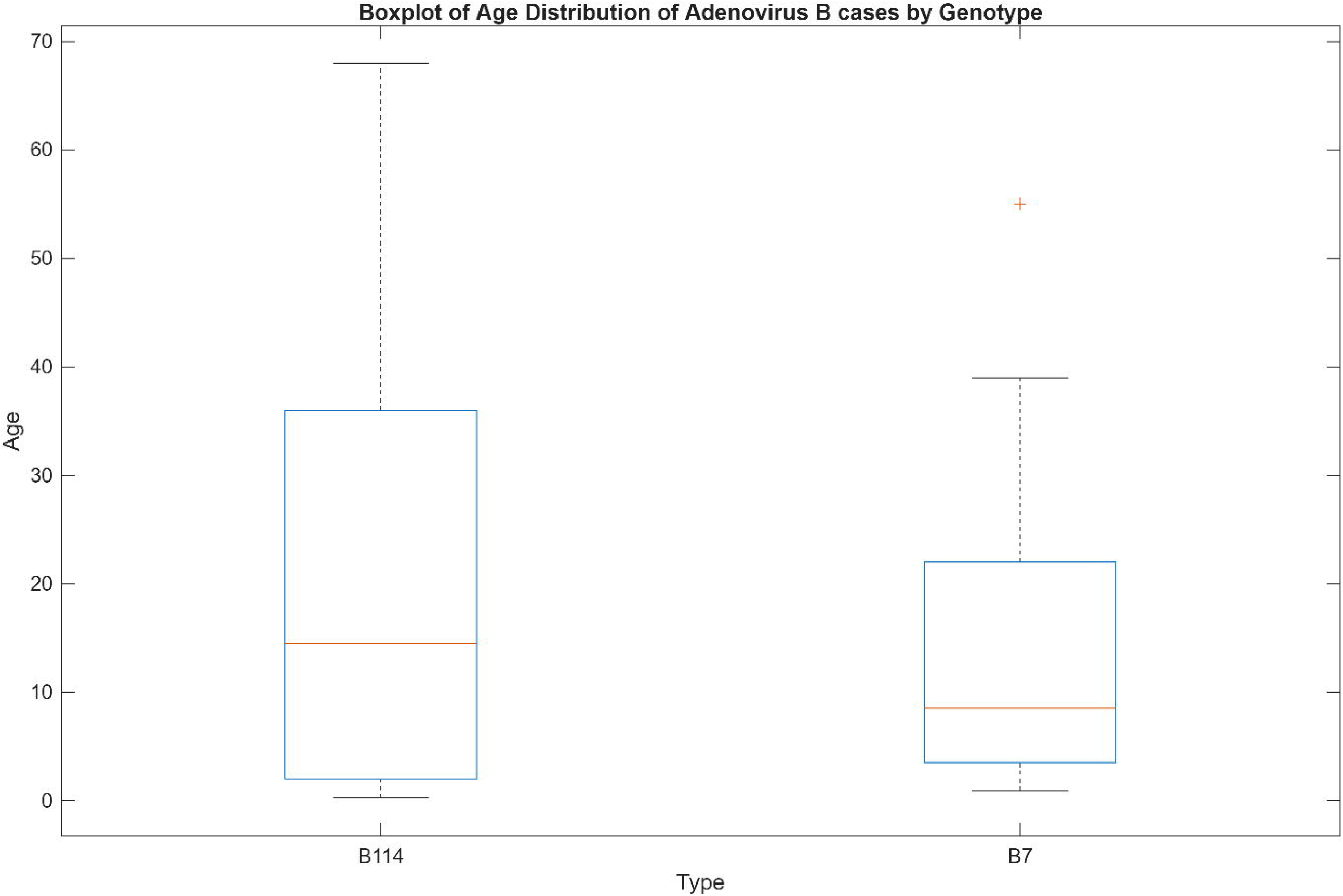
SimPlot++ analysis of similarities between B114 prototype sequence OR853835, HAdVB114_UKN22_23 (group 9) and HAdVB114_UKN22_7 (group 7). Group 7 is shown is pruple and group 9 is shown in green. The analysis was run with a window size of 200 bp and a step size of 20 bp.

Amino-acid alignments of the hexon and fibre genes were examined to determine whether any of these differences resulted in coding changes. No amino-acid differences were identified between the two representatives in the fibre gene. In the hexon gene, one amino-acid difference was identified at codon 204: the group 9 representative (HAdVB114_UKN22_23) carried aspartate (D), consistent with the B114 prototype sequence (OR853835.1), while the group 7 representative (HAdVB114_UKN22_7) carried asparagine (N). Using B114 prototype genome coordinates, codon 204 was mapped to hexon loop 1.

## Discussion

Until 2023, HAdV-B114 had not been recognised as a distinct genotype; samples carrying this recombinant genotype were routinely misclassified as HAdV-B3 because diagnostic typing based on partial hexon sequencing could not detect its B7-derived penton. Subsequent studies such as Ganzenmueller et al. performed WGS on samples from a large B3 outbreak in Germany in 2023, leading to the recognition that these genomes contained the recombinant capsid structure of B114 [5]. Retrospective BLAST analysis of the B114 prototype sequence then identified over 100 genomic sequences from the preceding two decades in GenBank, all previously labelled as HAdV-B3, with greater than 99.8% sequence identity to B114. We identify three previously published UK sequences from 2015 and 2016 (MW686785-7) as B114 [14]. In line with these findings, we conclude that B114 is a neglected, re-emerge t genotype rather than a novel one. The presence of B114 in our dataset before the pandemic is consistent with this interpretation, suggesting that the 2022-23 rise reflects post-restriction expansion of a pre-existing lineage rather than first emergence. The Nottingham dataset therefore provides the first UK context for interpreting the international B114 data reported from Germany, the USA, China and Japan [5, 16].

However, the rise in typed B114 genomes in our dataset should not be interpreted simply as reflecting population incidence, as changes in healthcare attendance, social mixing and ravel across the restriction period may have influenced detected case numbers independently of viral transmission [17]. Additionally, a larger susceptible population after a period of reduced exposure may have amplified local spread [18]. This dataset comprises specimens originating from symptomatic patients diagnosed in secondary care rather than systematic population surveillance, which supports a hypothesis of local pre-pandemic circulation of B114 but does not allow precise inference about underlying population incidence or the relative contribution of these mechanisms.

The identification of 24 genomically identical HAdV-B114 genomes collected over an eleven month period raises important questions about what consensus-level genomic identity can and cannot establish in the context of adenovirus surveillance. On the surface, a cluster of this size, sharing zero pairwise SNP differences might appear compatible with a single continuous transmission chain. However, the associated metadata argued against this interpretation. The cluster spanned eleven months, encompassed a broad case age range, included both ocular and respiratory specimens and the samples originated from multiple different clinics. The metadata also revealed small groups of cases within the large identical genome cluster that shared clinic source and collection dates with short windows. Taken together, the temporal dispersion, mixed sample types and multiple clinic origins of the cluster was more consistent with repeated detection of a widely circulating community strain than with sustained nosocomial spread, with the temporally and spatially close groups of sequences raising the possibility of occasional localised onward transmission within that broader background. However, the possibility of localised onward transmission cannot be confirmed without individual level epidemiological linkage data, which were not available for this study [14, 19–21]. The available metadata nonetheless provide evidence arguing against a single nosocomial transmission chain, illustrating a practical value of genomic surveillance: even when sequence data cannot confirm transmission, it can flag clusters of closely related genomes for prioritised epidemiological follow-up, helping to direct contact tracing or infection control investigation more efficiently than clinical observation alone.

The inability to confirm transmission patterns confidently from genomic data alone, however, reflects a broader challenge in adenovirus genomic surveillance. Distinguishing genuine nosocomial transmission from repeated importation into a clinical setting is crucial for infection-control decisions, but this is considerably more difficult for adenoviruses than for some other pathogens. For example, the relatively rapid accumulation of mutations in SARS-CoV-2 [19, 20, 22] during the COVID-19 pandemic meant that sequences from unrelated infections typically differed from one another, giving sequence data meaningful discriminatory power for inferring whether two infections were part of the same transmission chain. In contrast, adenoviruses accumulate sequence differences more slowly [23, 24], meaning that infections separated in time or source may remain genomically identical for extended periods, making consensus-level WGS insufficient alone to resolve individual transmission events.

The identification of several monophyletic groups within the Nottingham HAdV-B114 sequences raised the question of whether differences reflected a recent recombination event in a single genomic region. B114 groups 7 and 9 differed by 16 SNPs across the genome, representing a small degree of divergence overall, with no amino-acid differences identified in the fiber gene. The two lineages are not clearly separated in hexon and fiber gene trees (data not shown), highlighting the added value of WGS over single-gene typing for resolving fine-scale lineage structure within a genotype.

The only amino-acid difference identified between the two sub-lineages in these capsid genes was at hexon codon 204, located in loop 1; variation at this position has not been previously reported in HAdV-B114 or the closely related genome type HAdV-B3a. The hexon protein is the primary target of the host immune response, with serotype-specific neutralising antibody epitopes localised in seven hypervariable regions (HVRs) contained within two surface-exposed loops: loop 1 (HVR 1-6) and loop 2 (HVR 7) [25]. A difference at codon 204 is therefore of interest given its location in an antigenically exposed region. The change from aspartate to asparagine at this position also represents a loss of negative charge, which may be of functional relevance given the surface-exposed location of this residue, although this remains speculative without experimental validation.

In conclusion, this study demonstrates that WGS can define circulating adenovirus genotypes, reveal lineage structure, resolve fine-scale lineage structure and identify candidate sites of adaptive divergence within a clinical dataset. This provides the first detailed genomic characterisations of HAdV-B114 circulating in a UK clinical setting, addressing a gap in UK species B adenovirus genomic data.

## Data availability

Genome sequences are available on GenBank with accessions QB006051 - QB006118.

## Declarations of interest

CH’s lab receives unrelated funding through the BBSRC iCASE DTP, in collaboration with

Virothera Ltd.

---

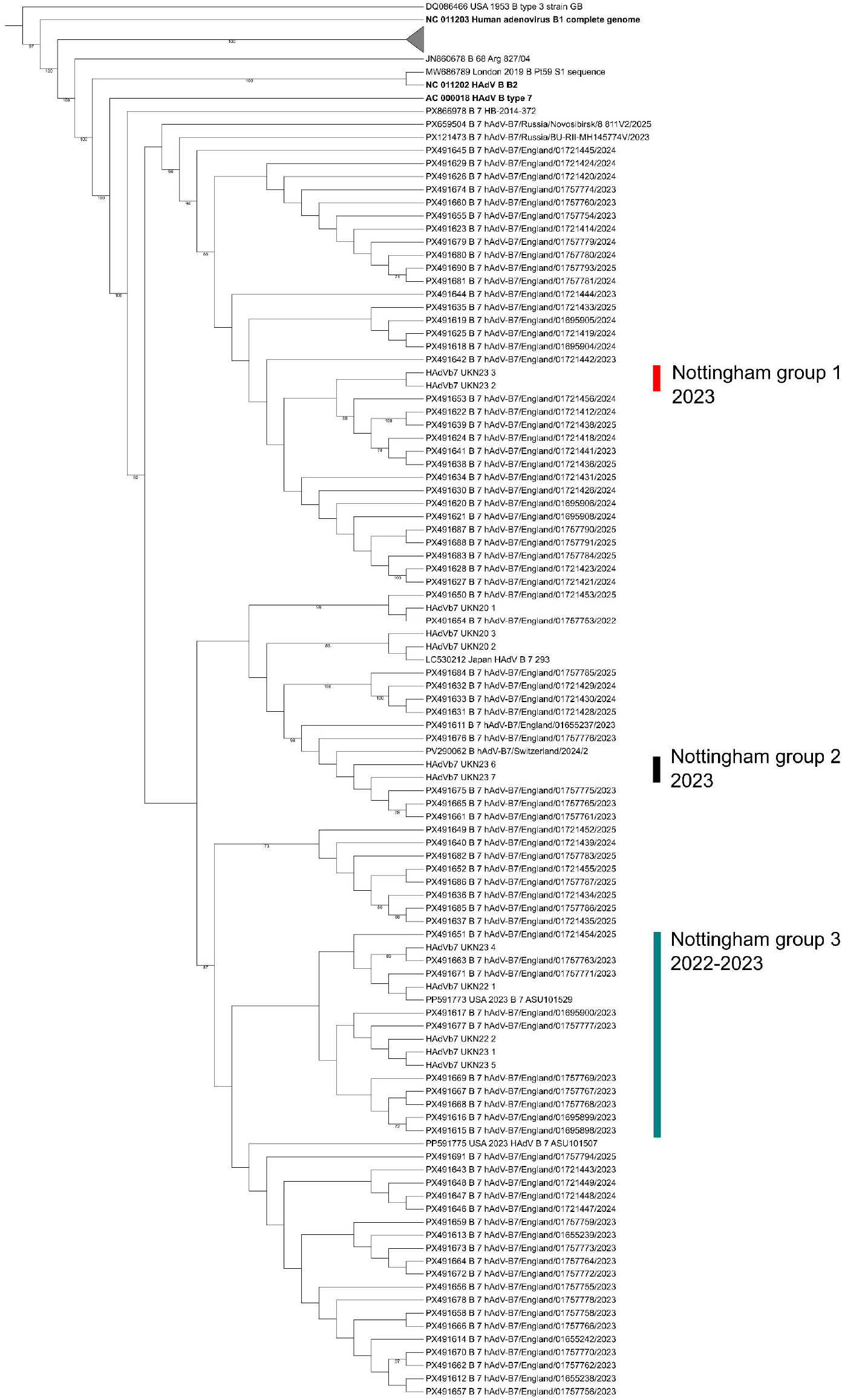

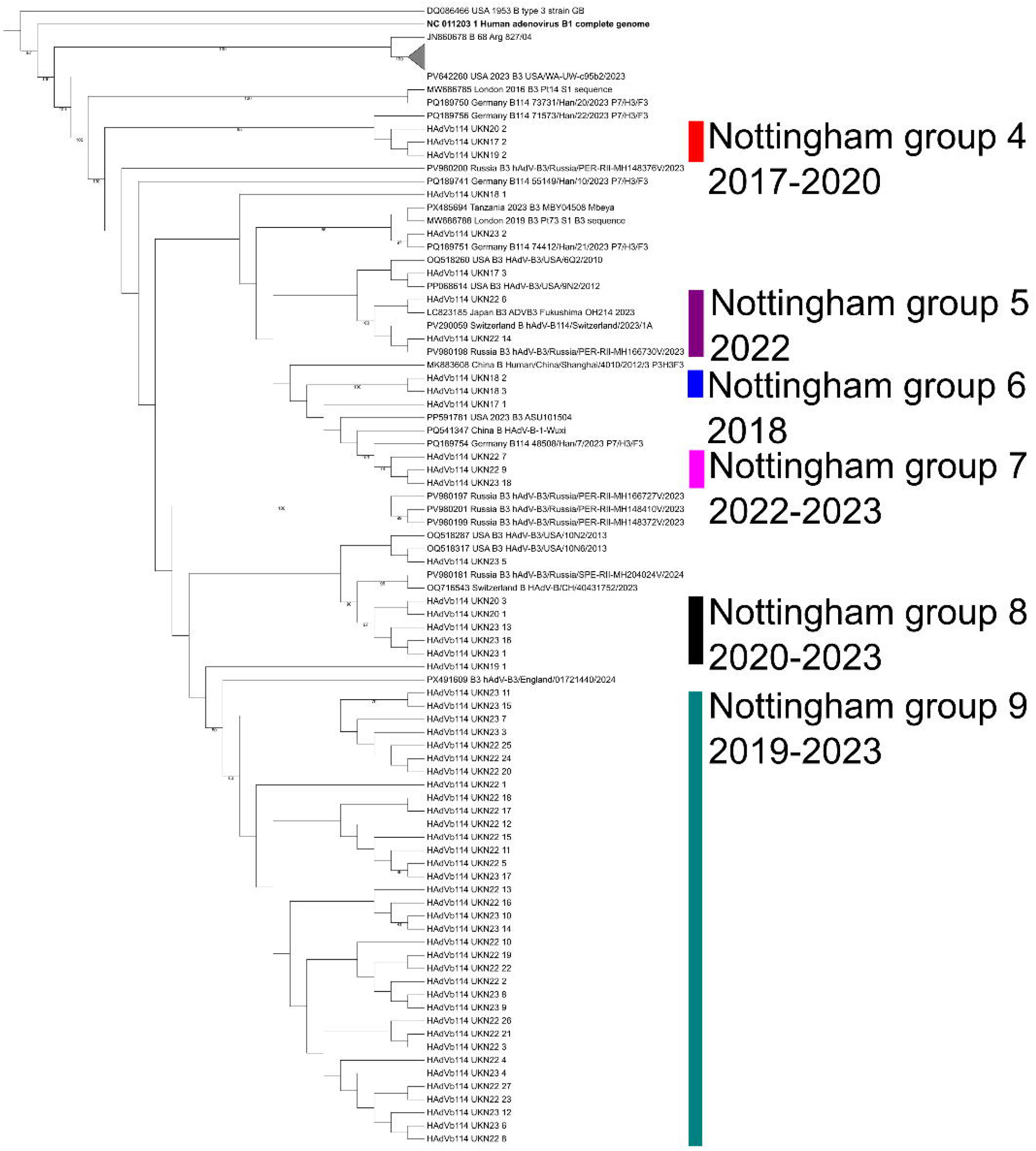

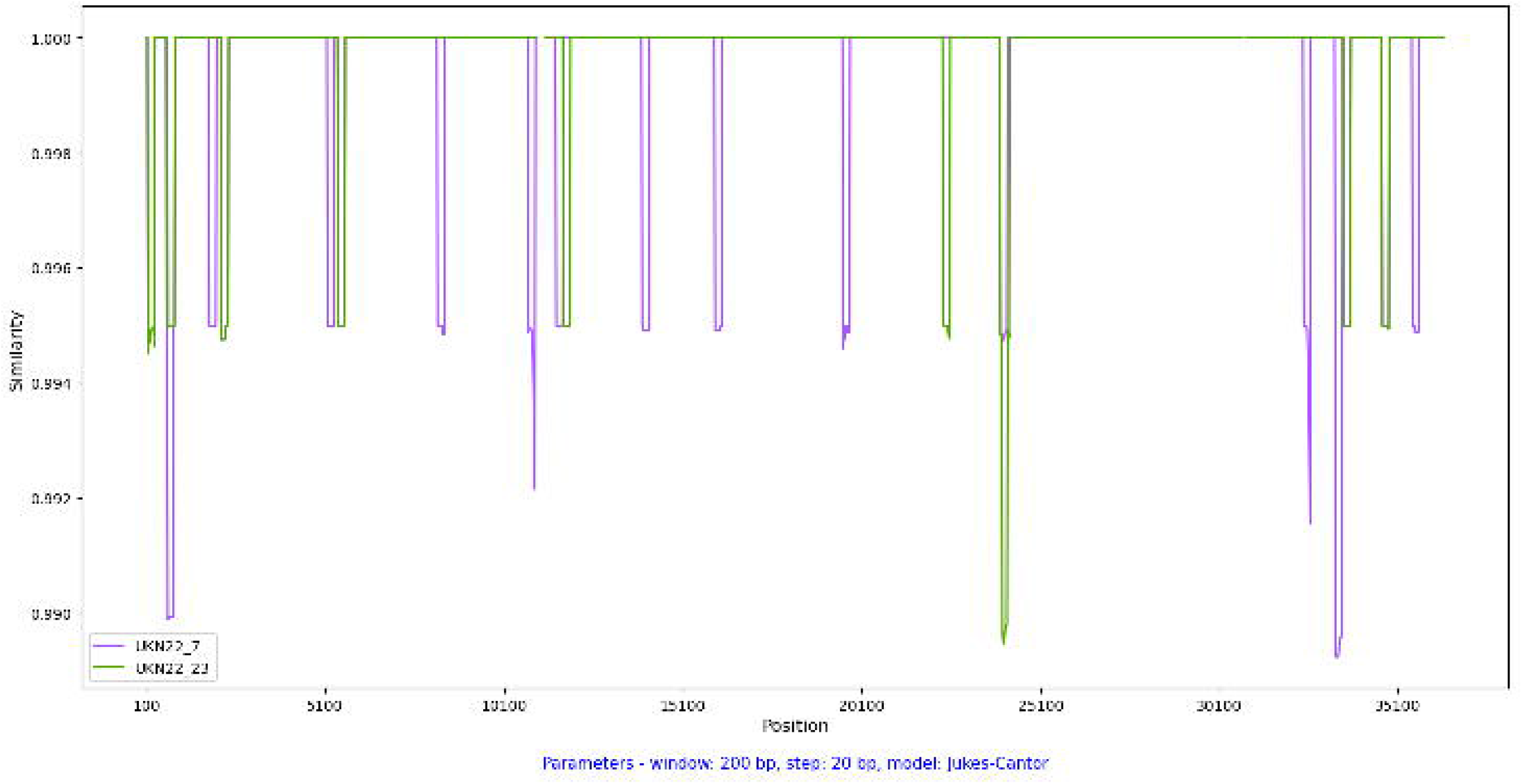

